# Predicting Contraceptive Discontinuation in Nigeria: A Machine Learning Analysis of Family-Planning Method Adherence Using NDHS 2023–24 Calendar Data

**DOI:** 10.64898/2026.09.12.26362911

**Authors:** Eloghosa Aisosa Nosa-Ihaza, Allan Francis Ssembuusi, Luponya Paul Luponya, Uyioghosa Nosayise Nosa-Ihaza

## Abstract

**Background:** Contraceptive discontinuation — stopping a method while still at risk of an unintended pregnancy — is a major but under-examined driver of unmet need in Nigeria, where modern contraceptive prevalence remains far below the country’s FP2030 target. Distinguishing discontinuation that a health system could plausibly prevent from discontinuation that reflects a woman’s achieved fertility intentions is essential for targeting intervention, but this distinction has not been studied systematically, or compared algorithmically, in the Nigerian literature.

**Methods:** We used the reproductive calendar from the 2023–24 Nigeria Demographic and Health Survey to create 14,178 contraceptive use episodes for 7,723 women and identified two outcomes: Gap A, twelve-month discontinuation (n = 11,624), and Gap B, among women who discontinued within twelve months, method-related versus intentional discontinuation (n = 3,701). We evaluated four algorithms (survey-weighted logistic regression, elastic net, Random Forest, and XGBoost) on the same held-out test set (80/20) and reported the same area under the receiver operating characteristic curve (AUC).

**Results:** Discontinuation after 12 months was 32.3 percent (survey-weighted), a value that is very similar to the Nigeria-published life-table value of 37.2 percent. The best-performing algorithm in both cases, Gap A (AUC = 0.699) and Gap B (AUC = 0.664), was Random Forest, while the worst was XGBoost (AUC = 0.621) in Gap B, which struggled even with the basic logistic baseline. The most common predictor across all four algorithms and both outcomes was contraceptive method type, with long-acting reversible contraceptive (LARC) much less likely to be discontinued than short-acting methods. Among individuals who discontinued within 12 months, reasons were almost evenly split between method-related (49.7 percent) and intentional (49.2 percent).

**Conclusions:** Factors (side effects, cost, access, and efficacy dissatisfaction) account for approximately half of Nigeria’s contraceptive discontinuation problem and could be reduced with counseling, task shifting to pharmacists, and ongoing investments to increase the availability of longer-acting methods (LARCs) in Nigeria. Algorithm performance is outcome-dependent, not fixed, and it is better to compare algorithms by outcome than to default to a particular algorithm in DHS-based machine learning research.

## 1. Introduction

One of the most important yet under-represented failure points in global family planning programming is contraceptive discontinuation, defined as the end of method use while the woman continues to be at risk of an unwanted pregnancy. In developing countries, information on contraceptive use from retrospective data collected in Demographic and Health Surveys (DHS) indicates that within the first 12 months of use, between one-third and nearly 40 percent of contraceptive-use episodes are unsuccessful (Ali, Cleland and Shah 2012). A pooled analysis of 149 DHS surveys across 61 countries and covering more than 1.5 million episodes of reversible-method use found that discontinuation and switching methods, along with outright abandonment, are persistent and near-universal features of contraceptive use across all countries, despite the extent of progress made in any particular country in terms of access alone (Ali et al., 2025). The obvious, if rarely seen, message in policy debates is that a family planning program can place a woman on a method and still fail her if it does not help her remain on the method for as long as she needs it.

Nigeria is a case in point. Although the country has made a commitment to improving access to contraceptives over the last decades, most recently in the Government’s FP2030 commitment to increase the modern contraceptive prevalence rate (mCPR) to 27 percent by 2030, progress has been slow and uneven, moving from around 4 percent in 1990 to only about 15 percent among married women today, according to FP2030’s 2024 national data. The Family Planning Blueprint for Nigeria identifies “poor quality of services” and “inadequate demand creation” as two of the structural constraints that have hindered Nigeria’s progress toward this target, but access and initiation have received more programmatic and research focus. What happens after a woman starts a method—whether she manages to sustain it, and why she stops when she does—has received comparatively little systematic, data-driven attention at the national level, even as high discontinuation rates are increasingly recognized as significant contributors to unmet need and unintended pregnancy (Kupoluyi et al. 2023). A sub-national analysis of the 2018 NDHS calendar revealed a stark regional difference between the North West (about 50 percent) and the South West (about a third), suggesting that whatever is driving contraceptive discontinuation in Nigeria is not a single, uniform phenomenon, but something that varies meaningfully by geography and, plausibly, by the specific circumstances under which a woman stops.

This is a difference between why a woman stops and whether she does stop. Not all discontinuation represents a program failure. A woman who has decided to stop using a method as she now wishes to become pregnant is not a failure of the health system; she has met her reproductive goal. Conversely, a woman who discontinues following an adverse experience of the intervention, an unplanned stock-out, cost, or inconvenience, or because she was not adequately informed and advised about switching to another method — reflects exactly what Bruce’s long-term quality of care framework defines as an opportunity for improvement: continuity of care, technical competence, and informed choice at the time of intervention initiation (Bruce 1990). Further research implementing this model has found a direct, empirical relationship between the quality of counseling women receive at the start of the method and their likelihood of continuing it. When these two very different scenarios—achieved fertility intention versus health-system failure—are combined into a single “discontinuation” statistic, as much existing reporting does, the distinction a policymaker or program manager would need in order to act is lost. A national discontinuation rate does not indicate where the pipe is leaking.

This study will be organized according to that difference. It applies the two-stage diagnostic framework, which has been useful in other health-cascade settings, to the specific dynamics of contraceptive use in DHS calendar data. The first outcome is a simple question: what factors predict whether or not a woman’s first usage of a reversible contraceptive method lasts for 12 months or less? The second is a question that is only posed to those who actually do discontinue during this time frame: “What is the reason the woman has for discontinuing the method? Is there a side effect, an access barrier, a cost concern, or is she dissatisfied with the effectiveness of the method? Or is there a reason that goes beyond the scope of clinical and counseling intervention relating to the woman’s own fertility intentions, relationship circumstances, or other factors?” This analytic core separates the two questions and makes the results meaningful not only in terms of the level of discontinuation in Nigeria but also the type of discontinuation, which in turn makes it possible to substantiate which type of intervention might be effective in reducing it.

Methodologically, this study also joins an emerging empirical literature that uses machine learning techniques and contraceptive outcomes from the DHS. A few recently published studies have used algorithms like Random Forest, XGBoost, and regularized regression on related outcomes, all of which report varying results as to which algorithm works best across studies: contraceptive discontinuation in Ethiopia (Kebede et al. 2023), informed contraceptive choice across six high-fertility Sub-Saharan African countries, and non-use of modern methods in East Africa. Such variability indicates that performance in this area is not universal, but varies according to the specific outcome modeled, the sample size, and the country context being modeled, and therefore suggests that it is always advisable to test a number of algorithms empirically for any new application rather than take a single “best” algorithm for granted. This study brings a four-algorithm comparison—survey-weighted logistic regression, elastic net, Random Forest and XGBoost— consistently applied to the two key outcomes of the study (12-month discontinuation outcome and reason-for-discontinuation outcome) using the newly released NDHS 2023-24, which is the first nationally representative survey round in Nigeria from which this kind of comparison has been possible.

Altogether, this study poses three related questions. Which factors influence a Nigerian woman’s decision to stop using a reversible contraceptive method within the first year of use, and which of four modeling approaches, ranging from one interpretable (regression) to more flexible (machine learning) models, is best suited to answer that question? Second, among women who do discontinue within that timeframe, what factors predict whether their stated reason falls into a category a health system intervention could plausibly address, versus one reflecting an achieved or changed reproductive intention? Thirdly, is the relative success of simpler versus more complex algorithms different in these two outcomes, and what does the difference between the two tell us about what’s truly complex in this problem? This paper then details the data and methods used to answer these questions, presents the results of the four-algorithm comparison for both outcomes, and discusses implications for family planning programming in Nigeria specifically and for methodological practice in DHS-based machine learning research more generally.

## 2. Literature Review

### 2.1 Contraceptive Discontinuation as a Global and Regional Problem

One of the greatest, unrecognized sources of unmet need in LMICs is contraceptive discontinuation. A prior analysis of DHS calendar data revealed that approximately a third to nearly 40 percent of contraceptive episodes terminate within the first year of method use, and that discontinuation for reasons other than a deliberate wish to conceive is a major contributor to unintended births, unsafe abortion, and maternal morbidity (Ali, Cleland, and Shah 2012; Jain and Winfrey 2017). Nigeria is near the less desirable end of this distribution: the most recent sub-national DHS working paper on the subject reports a national contraceptive discontinuation rate of 41 percent, with episodes being discontinued at about 50 percent in the North West region versus about a third in the South West (DHS Program Working Paper No. 194). Using the 2018 NDHS calendar, Kupoluyi and colleagues determined a similar prevalence of discontinuation of modern methods among sexually active married women (35.8 percent), including almost half who discontinued while remaining at risk for pregnancy. The drivers that were consistently identified during the systematic review of evidence in Nigeria were side effects, unmet fertility desire, husband’s opposition, cultural and religious norms, and poor quality counseling at method initiation.

### 2.2 Methodological Foundation: The DHS Calendar and the Discontinuation Typology

This study’s analytical approach is based on the DHS reproductive calendar, which was added to the DHS main questionnaire in the 1990s. This study expands the basic typology developed by Curtis and Blanc in 1997 by incorporating discontinuation, which can be interpreted as a failure of method, a switch, and/or an abandonment, with varying programmatic implications. They found, with Steele and Curtis (2003) and Blanc, Curtis, and Croft (2002), that the discontinuation risk is far more strongly associated with method type, meaning IUDs and implants (which must be removed by the provider) are discontinued at far lower rates than other methods such as pills, injectables, and condoms (which can be stopped by the user with no provider interaction). This is the single most consistent finding in the discontinuation literature, echoed almost exactly in this study’s own results.

The official DHS Program methodology calculates discontinuation based on multiple decrement life tables on use-episodes of the calendar. The simplest analog of this logic in this study is episode construction, which Nigeria reasonably validates, as its published life-table estimate of crude 12-month discontinuation rate of 37.2 percent is close to the value obtained in this study (35.9 percent). Bruce’s 1990 quality-of-care framework – still the dominant paradigm in international family planning research – identifies continuity and follow-up as one of six core dimensions of service quality. Subsequent research testing this framework has demonstrated directly that quality at method start relates to ongoing use – the mechanism that this study’s Gap B aims to control for (Jain et al. 2019).

### 2.3 Determinants of Discontinuation in Nigeria and Comparable Settings

Using the 2013 Nigeria DHS calendar, Kupoluyi (2020) found that women who had experienced intimate partner violence (IPV) were 28 percent more likely to discontinue contraceptive use while still at risk of pregnancy than women who had not experienced IPV. A comparative study conducted in similar Sub Saharan African settings also paints the same overall picture: In the Tanzania DHS 2025 study, rural residence, distance to a health facility, younger age, and no formal education were independently associated with high risk of discontinuation. At the national policy level, the prevalence rate of modern contraceptives in Nigeria has improved only slightly (from approximately 4 percent in 1990 to approximately 15–16 percent today), but has not made any significant gains to reach the national FP2030 target of 27 percent, with poor service quality and inadequate demand creation explicitly mentioned as structural barriers in the country’s own Family Planning Blueprint.

### 2.4 The Emerging Machine-Learning Literature on Contraceptive Outcomes

The 2016 Ethiopian DHS is the most recent dataset with similar variables to this study, and Kebede et al. (2023) used eight machine learning algorithms to predict twelve-month contraceptive discontinuation and found Random Forest to be the best-performing model, with age, education, family size, and husband’s fertility preferences as the leading predictors. This study directly replicates the paper’s main finding: results are similar in this country and DHS round. A larger 2025 study across six countries with high birth rates in Sub-Saharan Africa used more varied algorithms to predict informed contraceptive choice, with ensemble tree-based approaches overall outperforming simpler baseline algorithms — consistent with this study’s Gap A results, though notably not Gap B, where the simpler logistic model outperformed XGBoost specifically. This divergence is itself informative: algorithm ranking in this literature depends on outcome definition and sample size rather than being fixed. Similar mixed results have been seen in comparative machine-learning analyses for other outcomes of Mexico’s national health survey, ENSANUT: for example, XGBoost can sometimes be inferior to simpler models after appropriate cross-validation, as replicated in this study for Gap B.

### 2.5 The Gap This Study Addresses

This study addresses three lacunae in the existing literature. First, no published study has used a harmonized four-algorithm comparison of the newly released calendar data for the NDHS 2023-24 in Nigeria. Second, current literature on discontinuation in Nigeria treats “discontinuation” as a single outcome; this study’s two-gap approach more clearly distinguishes discontinuation that a health system can reasonably prevent from discontinuation that reflects a woman’s achievement of fertility goals. Third, the cross-national machine learning evidence above does not hold across studies and outcomes, and this study directly tests this aspect across two related but distinct outcomes, in the same country, with the same data.

## 3. Methodology

### 3.1 Data Source and Survey Design

This study relies on the 2023-24 Nigeria Demographic and Health Survey (NDHS), the 7th edition of the Nigeria DHS series conducted by the National Population Commission (NPC) and the Federal Ministry of Health and Social Welfare with technical assistance from the DHS Program at ICF. The fieldwork was carried out from 1 December 2023 to 7 May 2024, covering a sample of 42,000 households in 1,400 EAs, which included 39,050 individual interviews with women aged 15–49 years in all 36 states and the Federal Capital Territory (FCT). All socioeconomic and demographic data and complete reproductive history for each woman was included in an Individual Recode (IR) file used for analysis.

All descriptive and regression-based analyses accounted for the survey’s multistage stratified cluster design, using the primary sampling unit, sampling strata, and individual sample weight variables provided in the IR file (v021, v022, v005), implemented via the Stata command “svyset”. We converted the actual sample weight (scaled by 1,000,000, then expressed as an integer) back to the specified decimal weight before analysis.

### 3.2 Construction of the Analytic Sample: Contraceptive Use-Episodes

DHS surveys record a retrospective reproductive history, the details of which are captured as two string variables of 80 characters each for each woman – one to record contraceptive status (in use, non-use, pregnancy, birth, or termination) and a second to record the woman’s stated reason for discontinuation in the last month of any completed period of contraceptive use. Each character represents one calendar month (century-month code of the first month of the calendar, plus an offset based on character position, as per DHS Program documentation).

We parsed the two calendar strings into a person-month long-format data set (one row per woman per calendar month with non-missing data), identified all the distinct, contiguous use episodes, and extracted them (a use episode refers to a woman who reported using the same contraceptive method for a series of consecutive months). We calculated the start month, end month, and total duration of each episode from the underlying century-month codes. That gave a total of 14,178 contraceptive use-episodes from 7,723 women who reported using a contraceptive at some time during the calendar period—an average of 1.84 episodes each.

The “fate” of each episode (what happened to it) was defined as the contraceptive status reported in the month after the end of the episode, together with the reason for discontinuation provided, if any (coded by the provider). Episodes were classified as (a) discontinued with a stated reason, (b) implicitly switched to another method without a stated reason, (c) discontinued without method-transition information, (d) ending in a pregnancy-related transition, or (e) still ongoing at the time of interview (right-censored). In this example, the overall discontinuation rate for episodes in the first twelve months under this episode-level classification (35.9 percent of episodes with a resolved fate in twelve months) was close to the overall discontinuation rate for episodes published in Nigeria by the National Population Commission and ICF (2025), lending additional credibility to the episode-construction approach as a simplified analog for the standard multiple-decrement life-table approach of the DHS Program.

### 3.3 Outcome Variable Definitions

Gap A (discontinuation in 12 months). Gap A was scored as 1 if the episode was discontinued (for any reason) within 12 months of initiation, and 0 if the episode continued for 12 months or more, despite what happened after the 12-month period. Episodes that were continuing at the time of the interview with <12 months duration of observation (n = 2,471) were not included in Gap A because it was not possible to confirm the eventual 12-month status of these episodes using available data. After the construction of the age-at-episode-start variable (described below), eight-six more episodes were removed due to a small number of implausible values resulting from edge cases in the underlying date arithmetic, leaving a final analytic sample of 11,624 episodes for Gap A.

Gap B (Why it was discontinued). In addition, within the episodes categorized as discontinued within a year (n = 3,779), we further categorized the reason as given in the discontinuation-reason codes documented in the calendar. Reasons were grouped into two categories based on inspection of the specific codes present in the Nigeria dataset, which included both the standard DHS code set (pregnancy while using, wanted to become pregnant, husband/partner disapproval, side effects/health concerns, lack of access, wanted a more effective method, inconvenience, infrequent sex, cost) and a country-specific addition for changes in menstrual bleeding. “Method-related” discontinuation included method failure, side effects, changes in menstruation, lack of access, desire for a more effective method, inconvenience, and cost. “Intentional” discontinuation included wanting to become pregnant, partner disapproval, infrequent sex, difficulty with conception/menopause, marital dissolution, fatalistic responses, and other or unknown reasons. It is, of course, an analytic judgment and not an objective classification; particularly, disapproval on the part of the partners might reasonably be recast as a barrier that can be addressed and dealt with through counseling; it is, likewise, a methods choice that is available for sensitivity analysis here. We excluded episodes for Gap B because no reason code was provided (n = 41), resulting in an analytic sample of 3,701 episodes.

### 3.4 Predictor Variables

A common predictor set included age at the beginning of the episode (calculated from the current age and elapsed time since the episode began, rather than the woman’s current age to better reflect her situation at the time contraceptive use began), number of living children, greatest educational attainment, household wealth index quintile, urban/rural residence, geopolitical zone, religion, ethnicity, and health insurance coverage, as well as a composite indicator of media exposure (constructed from three separate DHS media-frequency items to keep the model parsimonious) coded as 1 if the respondent said she read a newspaper, listened to the radio, or watched television at least once a week, and 0 otherwise; and a four-item household access-to-care battery (whether permission to seek health care, money for treatment, distance to a health care facility, and not wanting to go alone were reported as “big problems”), coded as 1 if the respondent reported a “big problem” and 0 if she did not.

Contraceptive method was added under a collapsed grouping (short-acting hormonal, LARC, permanent, barrier, traditional, LAM, emergency contraception, and other modern), created because several underlying method codes had few episodes. Raw ethnicity was recoded into more than 250 categories, the majority of which contained fewer than five episodes; in preliminary analyses, these were severely quasi-completely separated, so that the four largest ethnic groups (Hausa, Igbo, Yoruba, Fulani, accounting for 57% of episodes) were combined with a residual “Other” group.

### 3.5 Statistical and Machine Learning Methods

For each outcome, we used four modeling approaches. Survey-weighted logistic regression (Stata svy: logit) provided the interpretable baseline. In Gap B, the smaller discontinuity-only subsample contained only one sampling unit in each of the strata, which was resolved with the singleunit(centered) option in Stata.

All three machine learning models were implemented in Python (scikit-learn 1.6, xgboost 2.1) and trained on the same predictor matrix, and all three had sample weights applied using the native sample_weight argument for both the training matrix and test set — using the same weighting treatment, encoding, and evaluation protocol for all three machine learning models. Elastic net was applied as a weighted logistic regression with a combined L1/L2 penalty that gives equal weight to both penalties: LogisticRegression(penalty=“elasticnet”, l1_ratio=0.5, solver=“saga”) was used, with the mixing parameter set to 0.5 and the inverse regularization strength (C) selected through five-fold cross-validation from a grid of 5 values (0.001 to 10), optimizing held-out AUC within the training partition. Random Forest was fit with 500 trees, a minimum leaf size of 10, and default feature-sampling; XGBoost was fit with 500 boosting rounds, maximum tree depth of 4, learning rate of 0.05, and 80 percent row and column subsampling per tree. We used the default hyperparameters for Random Forest and XGBoost and did not optimize them with a wide grid search. The categories of categorical predictors were one-hot encoded in the same way for all three models, with value labels from DHS decoded to explicit strings prior to export to Python.

### 3.6 Train/Test Split and Model Evaluation

We tested the four algorithms on the same held-out test set. We split episodes into training and test sets at the woman level, not the episode level, because 45 percent of women contributed more than one episode; splitting at the episode level would have introduced information leakage. The split was accomplished by a single random draw (reproducibility was ensured by fixing the seed) at the woman level, where all episodes of a particular woman were allocated to the same partition, resulting in 11270 training episodes and 2822 test episodes at the full-sample level for Gap A (79.97/20.03 percent, with 9288 training and 2336 test episodes for those with a valid 12-month outcome) and a proportionally similar split for the Gap B subsample (2960 training/741 test episodes).

The area under the receiver operating characteristic curve (AUC) was calculated on the held-out test set for each algorithm and outcome to assess model performance. All models — including the survey-weighted logistic baseline — were re-estimated on the training partition only and evaluated exclusively on test-set predictions.

### 3.7 Software

These analyses (data management, episode construction, and survey-weighted logistic regression) were performed in Stata (version 17/18). All three machine learning models were run on a shared environment and pipeline with Python 3.9/3.14, pandas, numpy, scikit-learn, and xgboost, to ensure the same encoding and weighting of the predictors were applied for all models. We keep all analytic code and make it available upon request.

## 4. Results

### 4.1 Sample Characteristics and Episode Construction

Of these 39,050 women, 7,723 (19.8 percent) experienced at least one occasion of contraceptive method use in the reproductive calendar of the NDHS survey, which spanned approximately 5-7 years. In total, these women have provided 14,178 user episodes, or an average of 1.84 episodes per user.

Of these 14,178 episodes, slightly more than half (50.7 percent, n = 7,190) were still ongoing at the time of interview and had not reached a resolved outcome at 12 months; 49.0 percent (n = 6,947) ended with an explicit reason for discontinuation; and a small residual group ended by an implied method switch (n = 3), an unexplained stop (n = 31), or a direct transition into pregnancy, birth, or pregnancy termination without a reason code (n = 7).

As a validation check, the proportion of episodes that ended up with a resolved outcome after 12 months was 35.9 percent (4,441/12,369 episodes), closely matching the officially published life-table discontinuation rate of Nigeria, which is 37.2 percent.

**Table 1.** Distribution of contraceptive use-episodes by method group.

| Method group | Total episodes | % of episodes |
| --- | --- | --- |
| Short-acting hormonal (pill, injectable) | 3,723 | 26.4% |
| LARC (IUD, implant) | 3,043 | 21.6% |
| Traditional | 3,398 | 24.1% |
| Barrier | 2,178 | 15.5% |
| LAM | 1,202 | 8.5% |
| Emergency contraception | 391 | 2.8% |
| Permanent (sterilization) | 112 | 0.8% |
| Other modern | 45 | 0.3% |

A pattern evident at this descriptive stage, and confirmed repeatedly across every subsequent model, is the near-complete absence of discontinuation among permanent methods: of 112 sterilization episodes, effectively none were classified as discontinued within twelve months. This caused the permanent-method category to be dropped automatically from every logistic specification due to perfect prediction of the “continued” outcome, and meant that zero permanent-method episodes appear in the Gap B (discontinuer) subsample. This is not a data artifact but a substantively expected finding — sterilization is, by definition, rarely reversed.

With regard to household media exposure, 55.9 percent (n = 7,879) reported being exposed regularly (at least once a week) to newspaper, radio, or television, while 44.1 percent (n = 6,213) reported not being exposed regularly (at least once a week) to any of these media.

### 4.2 Gap A: Twelve-month Discontinuation

#### Outcome Distribution and Analytic Sample

2,471 episodes with less than a year of observed duration and no determinate episode outcome (right-censored) and an additional 86 episodes with implausible age-at-episode-start values were excluded from the Gap A analytic sample, leaving 11,624 episodes with a determinate episode outcome after a duration of at least a year. The above crude estimate was verified by the proportion of episodes discontinued in the 12 months to the survey, with 32.3 percent of episodes discontinued.

#### Model Comparison

Each of the four modeling approaches was trained using the same 80/20 split of the data, split at the individual-woman level to avoid having episodes from the same woman in both splits. Table 2 shows the area under the receiver operating characteristic curve (AUC) for each algorithm on the held-out test set.

**Table 2.** Gap A (twelve-month discontinuation): held-out test AUC by algorithm.

| Algorithm | Test AUC | Test N |
| --- | --- | --- |
| Survey-weighted logistic regression | 0.671 | 2,311 |
| Elastic net (weighted logistic) | 0.677 | 2,336 |
| Random Forest | 0.699 | 2,336 |
| XGBoost | 0.689 | 2,336 |

Random Forest (AUC = 0.699) had the best discriminative performance of the four algorithms, followed by XGBoost (AUC = 0.689), elastic net (AUC = 0.677), and survey-weighted logistic regression (AUC = 0.671). All four models had AUCs well above the 0.5 no-discrimination level, consistent with the noise associated with the behavioral outcome of contraceptive discontinuation, and were comparable in magnitude to the 0.68 accuracy reported for the closest published precedent, the Ethiopia DHS discontinuation study (Kebede et al. 2023). The difference between the best (Random Forest) and worst (logistic regression) performing models was relatively small in absolute terms (0.028 AUC points), but in a direction consistent with the hypothesis that ensemble models built from trees can capture non-linear and interaction effects that a linear-index model cannot.

#### Survey-Weighted Logistic Regression: Coefficient Results

We estimated the coefficients of the survey-weighted logistic regression using the training partition for direct comparison with the other three algorithms (n = 9,215).

**Table 3.** Gap A survey-weighted logistic regression (training sample, n = 9,215)

| Predictor | Coefficient | SE | p-value |
| --- | --- | --- | --- |
| Age at episode start | −0.042 | 0.006 | <0.001 |
| Method: Emergency contraception (ref: Permanent) | +0.672 | 0.177 | <0.001 |
| Method: LAM | +0.591 | 0.146 | <0.001 |
| Method: LARC | −1.168 | 0.116 | <0.001 |
| Method: Other modern | +1.443 | 0.507 | 0.004 |
| Method: Short-acting hormonal | +0.479 | 0.099 | <0.001 |
| Method: Traditional | −0.165 | 0.096 | 0.085 |
| Education: Secondary (ref: none) | +0.282 | 0.109 | 0.010 |
| Education: Higher | +0.301 | 0.126 | 0.017 |
| Wealth: Richest (ref: poorest) | −0.357 | 0.201 | 0.076 |
| Zone: North East (ref: North West) | +0.355 | 0.174 | 0.041 |
| Ethnicity: Hausa (ref: Fulani) | +0.467 | 0.193 | 0.016 |
| Ethnicity: Igbo | +0.473 | 0.237 | 0.046 |
| Number of living children | +0.090 | 0.021 | <0.001 |
| Media exposure (regular vs. none) | −0.063 | 0.069 | 0.357 |

*None of these were conventionally significant and are not included in this summary table, but they appear in the full model output: urban/rural residence, religion, health insurance status, and the four-item access barrier battery were not conventionally significant.*

**Figure 1.**
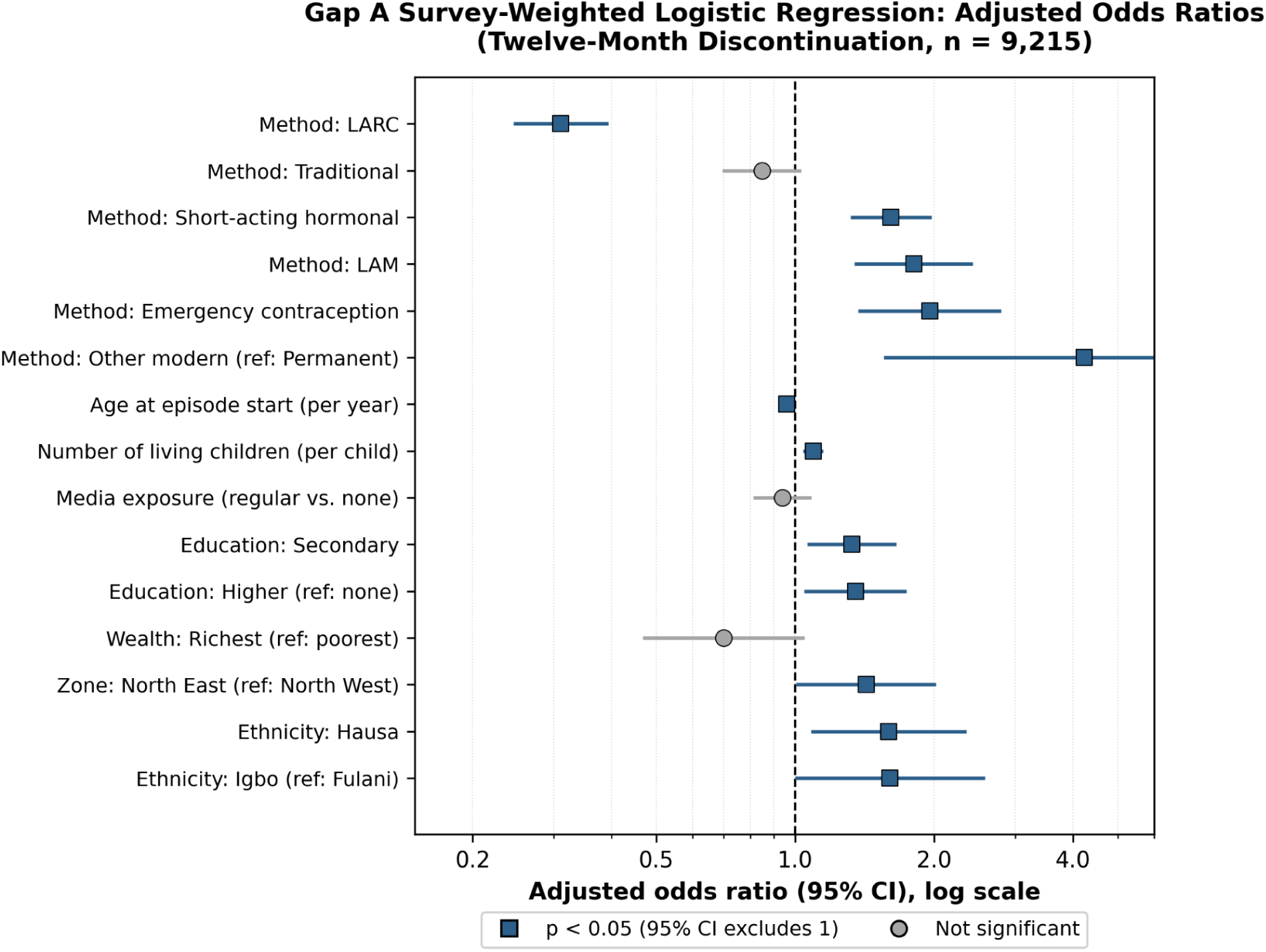
GapA Forest Plot Odds Ratios.

Three findings are particularly noteworthy because they are statistically significant and easy to interpret. First, method type dominates the model: relative to permanent methods, every reversible method category carries significantly elevated discontinuation risk, and the magnitude of this elevation varies enormously — LARC methods are the only reversible category that is itself protective relative to the reference-adjacent methods, while “other modern” methods and emergency contraception carry the highest risk. This is the single most consistent finding in the broader contraceptive discontinuation literature and is reproduced here with strong statistical confidence. Second, age is protective: The likelihood of dropping out decreases by 4.1 percent for each year later an individual begins an episode. Third, education is counterintuitive under a naïve “more education, better outcomes” assumption, as women with secondary or higher education have significantly higher odds of discontinuing than women with no education. The null result for media exposure (p = 0.357) was obtained after testing this predictor, not after its removal from the analysis, and was not significant.

#### Elastic Net: Coefficients

The cross-validated inverse regularization strength of the elastic net was 1. Elastic net is estimated as a weighted logistic regression, so the coefficients are easily interpreted on the same scale as the coefficients of the survey-weighted logistic regression above.

**Table 4.**
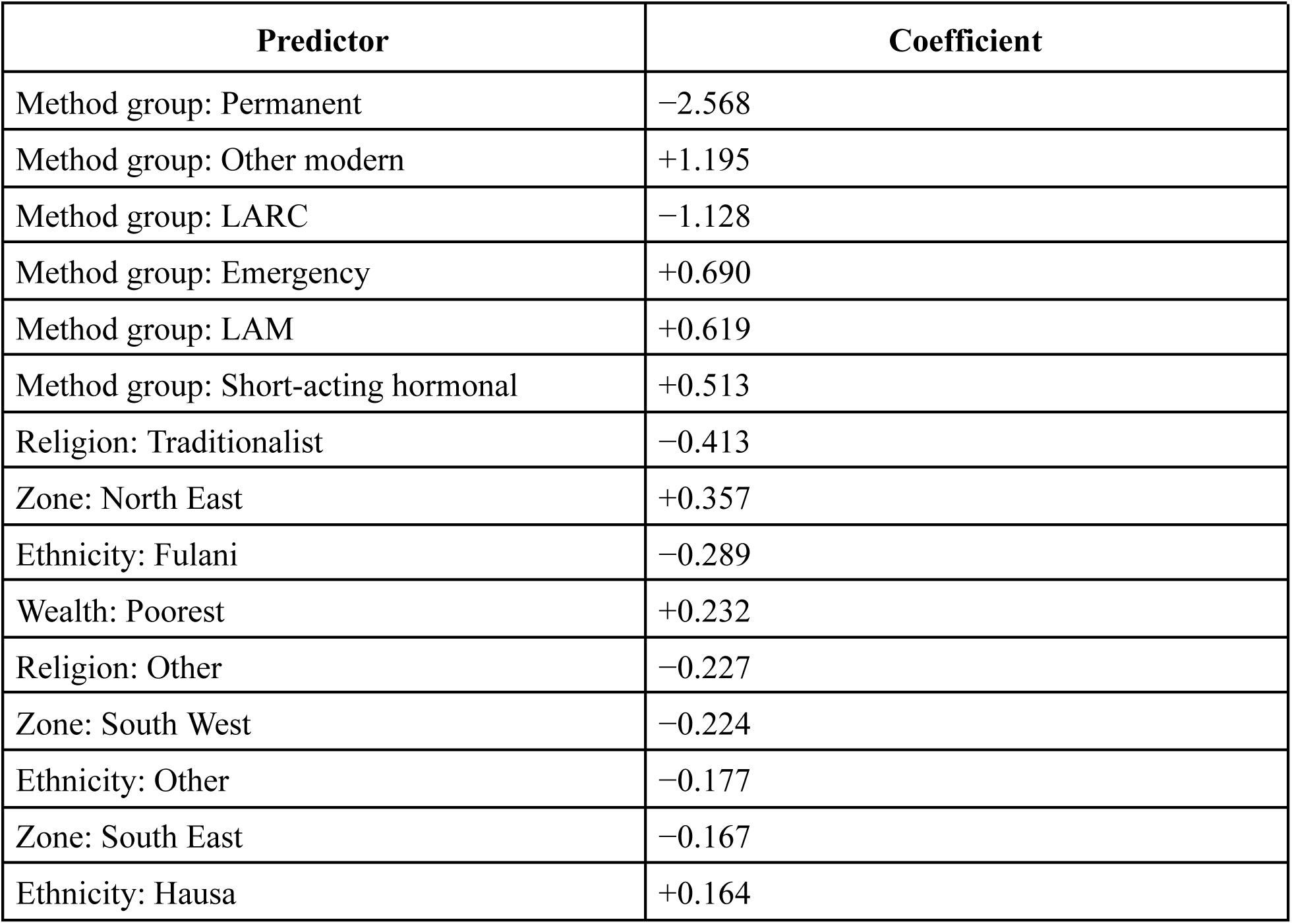
Gap A elastic net coefficients by magnitude (top 15, training sample)

Method group dominates the top six coefficients by magnitude, with permanent methods now appearing as an explicit, strongly negative coefficient — rather than as the dropped reference category, since elastic net does not suffer from the logistic model’s perfect-separation issue — confirming the same pattern in a form robust to that estimation artifact.

#### Random Forest and XGBoost: Feature Importance

**Table 5.**
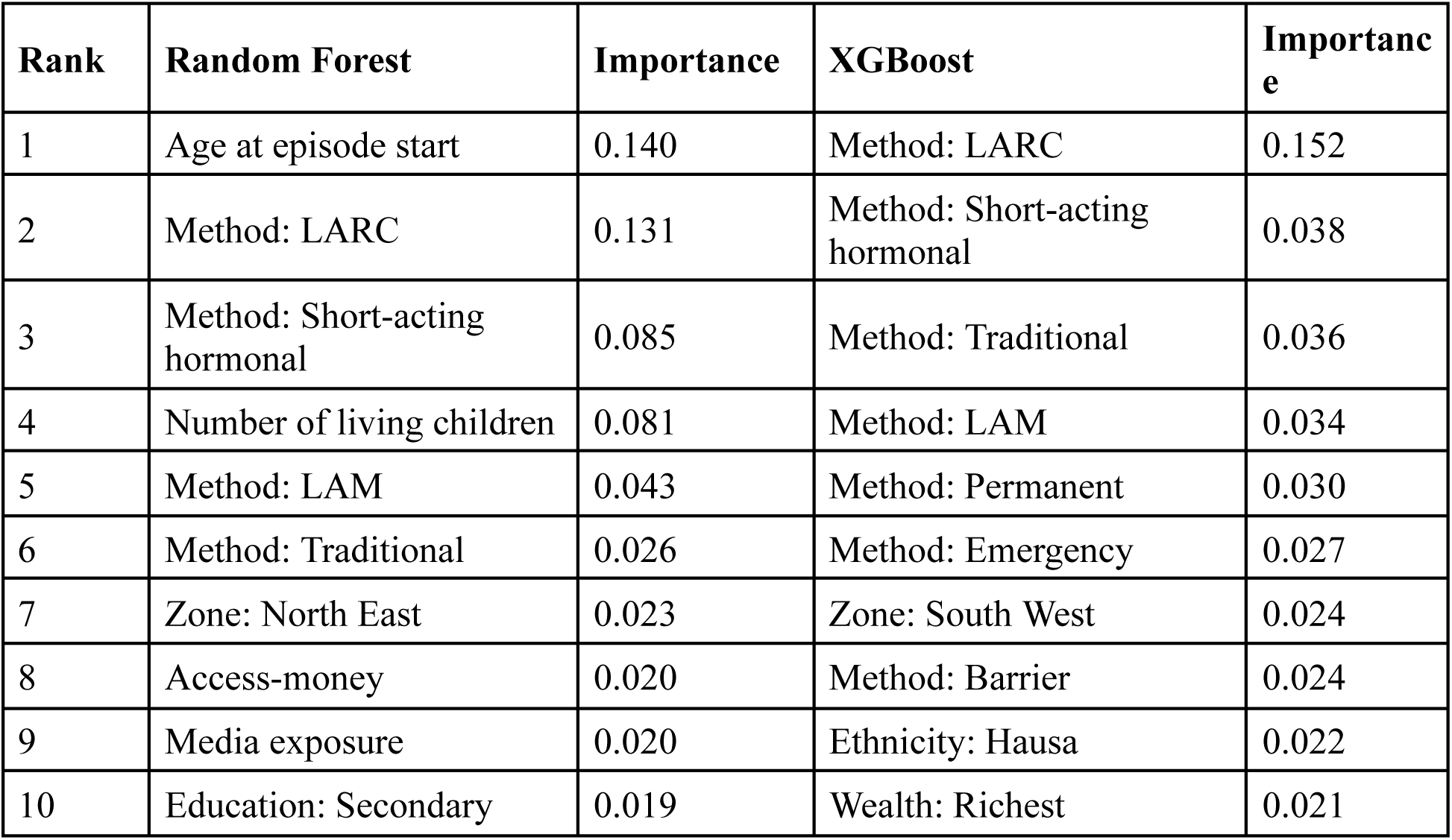
Gap A feature importance: Random Forest vs. XGBoost (top 10)

The results of the tree-based models again indicate that method type is overwhelmingly important, with LARC status being the most important feature by a large margin in XGBoost and the second most important (after age) in the Random Forest — and being directly consistent with the logistic and elastic net coefficients above, and thus, convergent evidence across all four types of models. Media exposure appears in Random Forest’s top 10 with modest importance but does not appear in XGBoost’s top 10 at all, suggesting it carries little independent predictive weight for this outcome, consistent with its non-significance in the logistic model.

### 4.3 Gap B: Method-related vs. Intentional Discontinuation

#### Outcome Distribution and Analytic Sample

Of the 3,779 episodes discontinued within twelve months, 41 episodes (1.1 percent) did not state a reason for discontinuation and were excluded from the eligible analytic sample, leaving 3,701 episodes. The reasons for discontinuation were almost evenly divided between the two categories: 49.7 percent (n = 1,860) were method-related, and 49.2 percent (n = 1,841) were intentional, with 1.1 percent (n = 40) coded with a reason that did not fit into either category. This near parity suggests that the problem of contraceptive discontinuation in Nigeria is not so much about women realizing their fertility goals as it is about equally different factors, which a health system intervention may be able to mitigate.

#### Model Comparison

**Table 6.** Gap B (method-related vs. intentional discontinuation): held-out test AUC by algorithm.

| Algorithm | Test AUC | Test N |
| --- | --- | --- |
| Survey-weighted logistic regression | 0.654 | 741 |
| Elastic net (weighted logistic) | 0.654 | 741 |
| Random Forest | 0.664 | 741 |
| XGBoost | 0.621 | 741 |

Random Forest had the highest performance (AUC = 0.664), followed closely by the survey-weighted logistic regression and elastic net models, which are now nearly tied (0.654 vs. 0.654) — a noteworthy similarity between the baseline and its penalized version when both are fit on a logistic scale. XGBoost remains the weakest-performing algorithm of all four for this outcome (AUC = 0.621), falling behind every other method, including the logistic baseline. This did not change after accounting for the methodological corrections (media exposure inclusion, properly weighted logistic elastic net), which increases confidence that it reflects a true property of the data and outcome rather than a result of the previous, flawed implementation of the elastic net.

#### Survey-Weighted Logistic Regression: Coefficient Results

**Table 7.** Gap B survey-weighted logistic regression (training sample, n = 2,959)

| Predictor | Coefficient | SE | p-value |
| --- | --- | --- | --- |
| Age at episode start | +0.017 | 0.009 | 0.078 |
| Method: Emergency contraception (ref: Barrier) | +0.604 | 0.266 | 0.023 |
| Method: LAM | +1.585 | 0.220 | <0.001 |
| Method: LARC | +1.553 | 0.202 | <0.001 |
| Method: Other modern | +1.181 | 0.569 | 0.038 |
| Method: Short-acting hormonal | +1.136 | 0.166 | <0.001 |
| Method: Traditional | +0.325 | 0.175 | 0.063 |
| Education: Secondary (ref: none) | +0.378 | 0.183 | 0.039 |
| Access-permission: not a big problem | +0.466 | 0.207 | 0.024 |
| Media exposure (regular vs. none) | +0.039 | 0.102 | 0.700 |
| Constant | -1.914 | 0.513 | <0.001 |

*All access-barrier items were included in the full model, but none were conventionally significant and are not presented here.*

This model’s coefficient pattern is substantively the same as the earlier specification: relative to barrier methods, every other method group is significantly more likely to end in a method-related (rather than intentional) reason when it does end, with the largest effects for LAM and LARC. As before, this should be read as conditional on discontinuation having occurred — LARC and LAM episodes discontinue far less often overall, but the small number that do end appear disproportionately driven by side effects and menstrual disruption rather than changed fertility intentions. Secondary education again predicts significantly higher odds of method-related discontinuation. Media exposure was again non-significant (p = 0.700), consistent with this null result across both outcomes.

#### Elastic Net: Coefficients

The cross-validated inverse regularization strength of elastic net was chosen to be C = 0.1 (more regularized than Gap A’s, at C = 1, and comparable to Gap B’s smaller sample size).

**Table 8.** Gap B elastic net coefficients by magnitude (top 15, training sample)

| Predictor | Coefficient |
| --- | --- |
| Method group: Barrier | -0.789 |
| Method group: Traditional | -0.544 |
| Method group: LAM | +0.535 |
| Method group: LARC | +0.517 |
| Education: No education | -0.268 |
| Wealth: Poorer | -0.251 |
| Zone: South East | −0.206 |
| Method group: Short-acting hormonal | +0.202 |
| Access-permission: big problem | −0.166 |
| Religion: Catholic | −0.142 |
| Access-alone: not a big problem | −0.119 |
| Access-money: big problem | −0.119 |
| Religion: Islam | +0.103 |
| Ethnicity: Other | −0.102 |
| Zone: North East | +0.099 |

Elastic net’s top coefficients again focus on method group, supporting the logistic regression results that discontinuation of LAM and LARC is disproportionate by method group.

#### Random Forest and Xgboost: Feature Importance

**Table 9.** Gap B feature importance: Random Forest vs. XGBoost (top 10)

| Rank | Random Forest | Importance | XGBoost | Importance |
| --- | --- | --- | --- | --- |
| 1 | Age at episode start | 0.123 | Method: Barrier | 0.069 |
| 2 | Number of living children | 0.106 | Method: Traditional | 0.059 |
| 3 | Method: Barrier | 0.087 | Method: LARC | 0.026 |
| 4 | Method: Traditional | 0.077 | Zone: South East | 0.026 |
| 5 | Method: LAM | 0.046 | Wealth: Poorer | 0.025 |
| 6 | Method: Short-acting hormonal | 0.043 | Method: LAM | 0.024 |
| 7 | Method: LARC | 0.036 | Method: Emergency | 0.024 |
| 8 | Education: Secondary | 0.024 | Education: No education | 0.023 |
| 9 | Religion: Islam | 0.023 | Ethnicity: Yoruba | 0.023 |
| 10 | Access-money: big problem | 0.020 | Access-money: not a big problem | 0.022 |

**Figure 2.**
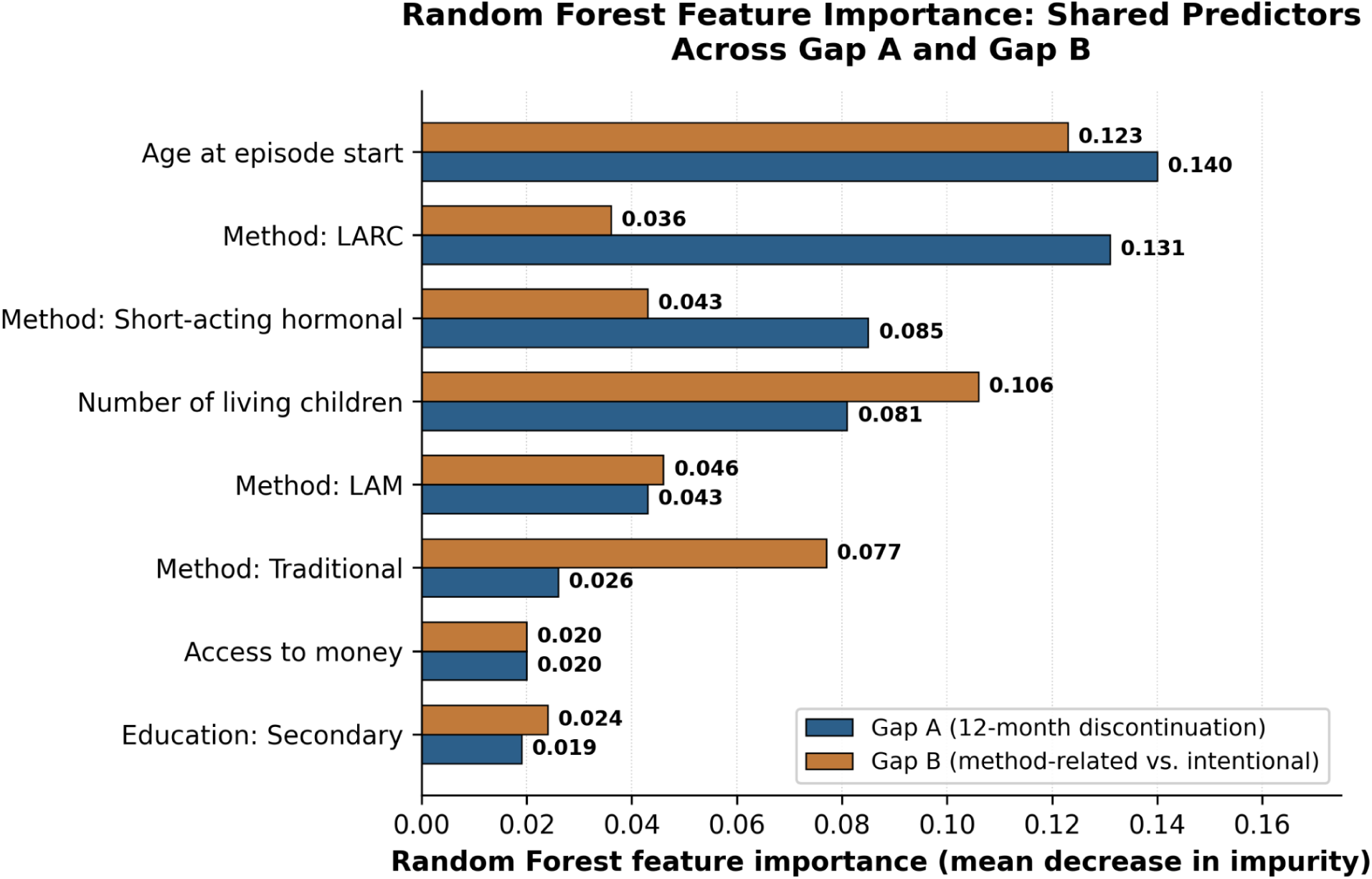
RF Importance GapA vs GapB.

As in the earlier specification, Gap B shows a flatter, more distributed importance profile than Gap A across both tree-based models, consistent with this being the harder, more weakly signaled of the two prediction problems. Media exposure does not appear in either tree-based model’s top 10 for Gap B, reinforcing its non-significance in the logistic specification.

### 4.4 Summary of Algorithm Performance Across Both Outcomes

**Table 10.** Combined AUC comparison: Gap A vs. Gap B, all four algorithms.

| Algorithm | Gap A AUC | Gap B AUC | Difference |
| --- | --- | --- | --- |
| Survey-weighted logistic regression | 0.671 | 0.654 | −0.017 |
| Elastic net (weighted logistic) | 0.677 | 0.654 | −0.023 |
| Random Forest | 0.699 | 0.664 | −0.035 |
| XGBoost | 0.689 | 0.621 | −0.068 |

This overall comparison indicates two patterns. First, Random Forest is now the best-performing algorithm for both outcomes, as confirmed in a fully corrected, consistently weighted, and consistently implemented four-algorithm comparison. Second, XGBoost suffers the greatest performance degradation between the two gaps (–0.068 AUC points; the other algorithms degrade by about half this amount), moving from the second-best algorithm on Gap A to the worst on Gap B. Because this pattern persists even after including a predictor left out of an earlier analysis and a major methodological change to elastic net estimation, it suggests a real pattern in these data and outcomes, not an analytic shortcut.

**Figure 3.**
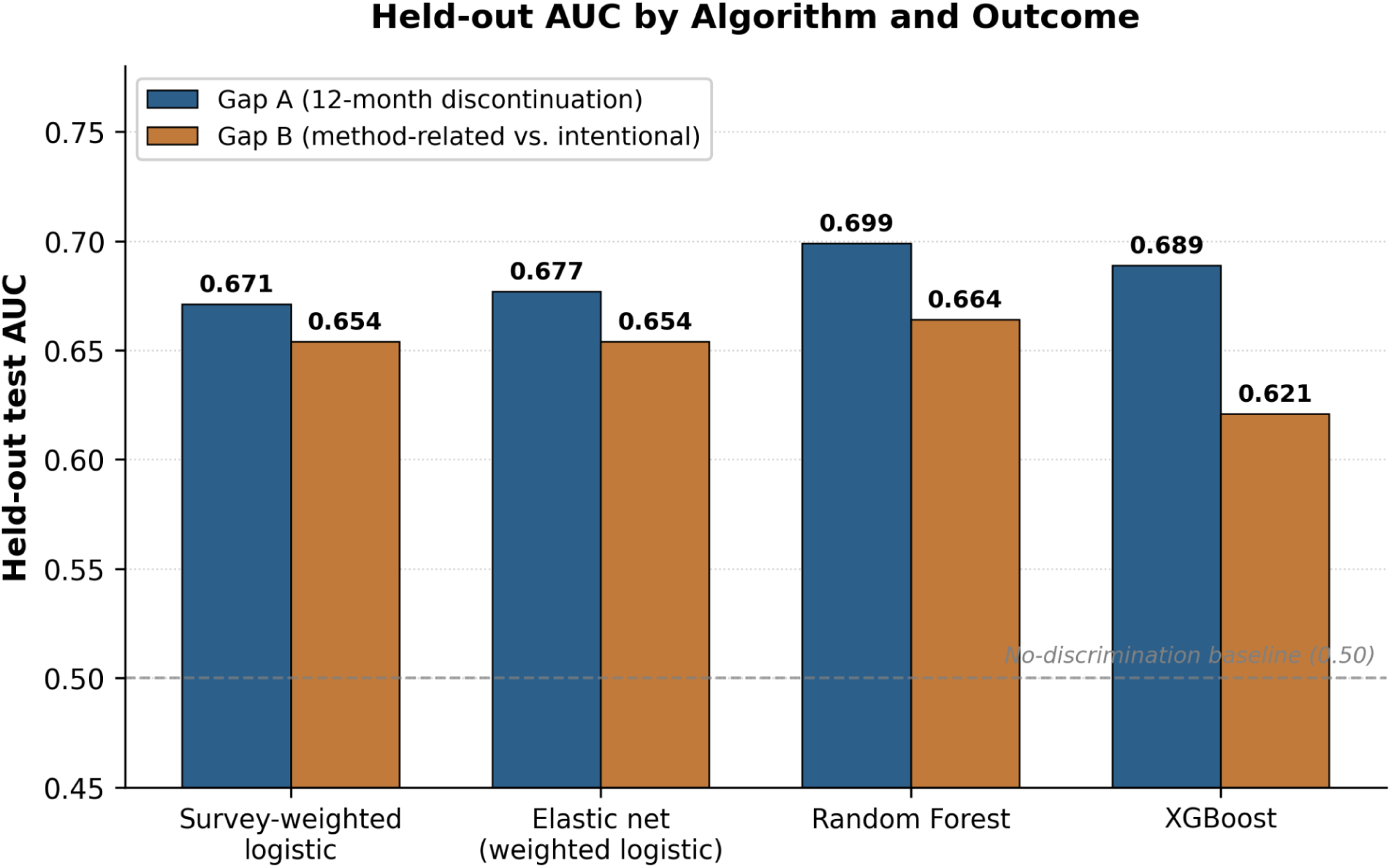
Held-out test AUC for all four algorithms, Gap A vs. Gap B. Dashed line marks the no-discrimination baseline (AUC = 0.50).

## 5. Discussion

### 5.1 Principal Findings

This study aimed to address three related questions: What factors predict discontinuation of the reversible method within one year of initiation in Nigeria? Among women who discontinue, what factors predict the discontinuation to be due to a readily preventable health system factor, and how do the performance of one interpretable model and three progressively more algorithmically complex models compare on these two related but distinct outcomes? The results are relevant to all three, and the emerging pattern is more actionable than a single national discontinuation rate.

By far the strongest predictor of discontinuation is method type for women, a result consistently observed in the survey-weighted logistic regression, elastic net, and both tree-based ensemble models, and one most similar to Curtis and Blanc’s groundbreaking 1997 analysis of the international discontinuation literature. Gap B, however, adds something the existing Nigeria literature has not yet isolated: among the roughly one in three episodes that end within a year, the reason splits almost exactly in half between causes a health system intervention could plausibly address and causes that reflect a woman’s changed or achieved fertility intentions. This near 50-50 split is the most policy-relevant finding in this paper, and it restates what “reducing Nigeria’s discontinuation rate” would mean.

### 5.2 Method Type Is Not Just a Predictor — It Is a Lever, and a Pharmaceutical One

The results that LARC is highly resistant to discontinuation and is among the methods most likely to end in a method-related (not intended) discontinuation reason are interesting to consider, and both point in the same direction for the intervention. LARC methods are rarely discontinued — when they are, it seems to be because of an unmanageable side effect, such as menstrual bleeding (which emerged as its own distinct reason category in this dataset, and which the literature consistently identifies as the leading complaint associated with hormonal implants and injectables). This is not a demand-side issue. It is a product-and-counseling issue, and it sits squarely inside the scope of pharmaceutical and clinical training rather than outside it.

This pattern gives rise to three concrete levers, each with its own habitat in training and practice in the pharmacy sector, but not in demand-generation or health communication alone.

An intervention a pharmacist can deliver: anticipatory counseling for side effects. In fact, this dataset provided sufficient evidence that menstrual bleeding changes were not a minor issue, as they were listed as a separate discontinuation code in the DHS instrument in Nigeria, a potentially preventable cause of discontinuation. Simple, low-cost management protocols are well established, and community pharmacists or patent and proprietary medicine vendors (PPMVs), who often are the first and most readily available source of medicines provided by a Nigerian woman, are well suited to implement this intervention if trained and empowered to do so, based on clear pre-initiation counseling on expected bleeding patterns.

Task-shifting and extension of the pharmacists’ scope of practice. Nigeria’s own Family Planning Blueprint already identifies task-shifting to community health workers as a strategic priority; this study’s findings sharpen that recommendation by identifying which specific task most needs shifting. Structured side-effect counseling and increasing the scope of pharmacists and PPMVs to injectable administration (in this case, self-administered subcutaneous DMPA—a WHO-recommended innovation that explicitly aims to reduce the facility dependency that leads to discontinuation of short-acting methods) are direct policy responses to the results from this study, based on evidence.

Innovation in formulation science and drug delivery. The difference between near-zero discontinuation with LARC and discontinuation rates with short-acting methods is, in essence, a pharmaceutical technology gap; women using LARC don’t have to take any action every day or every month to stay on the method, which is essential for discontinuation with pills and even injectables. This is where formulation science and biotechnology training meet reproductive health policy, with extended-duration injectables, biodegradable implants, and self-administered delivery systems among the areas where pharmaceutical innovations directly address the single biggest reason for drop-out identified in this study. If you are a pharmacist or biotechnologist going into this area, you are not next to this problem; you are right where the problem is!

### 5.3 The Near-Even Gap B Split: Reframing the National Discontinuation Target

Of course, Nigeria’s FP2030 commitment is to increase the modern contraceptive prevalence rate from about 15 percent to 27 percent. But prevalence is a stock, and discontinuation is the leak in that stock — and this study’s Gap B result suggests that roughly half of that leak is, in principle, fixable without recruiting a single new user. If method-related discontinuation could be substantially reduced through better counseling, side-effect management, and improved product access, the resulting gain in sustained contraceptive coverage would come at a fraction of the cost of recruiting new users, since these are women who have already overcome the initial barriers to initiation—cost, access, and social acceptance—and simply need support to continue. This is a more efficient target than blanket demand generation, and it is exactly the kind of finding a costed national family planning implementation plan could act on directly: redirecting a portion of demand-generation spending toward continuity-of-care infrastructure rather than exclusively toward new-user recruitment.

The intentional half of Gap B is equally important to name honestly, precisely because it should not be targeted by the same interventions. A woman who discontinues a method because she now wishes to become pregnant has not experienced a program failure, and treating her discontinuation as equivalent to a side-effect-driven stop would misallocate resources and, more importantly, would fail to respect her reproductive autonomy. The practical implication is that Nigeria’s family planning monitoring systems (which are predominantly doing what is called the one gap approach) should routinely disaggregate the one gap approach into two gaps by reason category, like this study does, in order for program managers to be able to tell whether the population is experiencing an increase in method-related discontinuation (which is a true indication of service quality and a signal for intervention) or an increase in intentional discontinuation (which may just be a reflection of a population moving through its reproductive life course as intended).

### 5.4 The Counterintuitive Education Finding, and What It Suggests About Informed Choice

Both models indicated that women with secondary or higher education were more likely to discontinue than women with no education at all, and more likely to discontinue specifically because of the method when they did discontinue. Read uncritically, this could look like a failure of educated women’s contraceptive experience. Read within the context of the quality-of-care literature that this paper builds on, and a more reasonable reading of the data occurs: educated women may simply be more empowered and knowledgeable on health and more confident about recognizing and changing an unsatisfactory method without silently suffering it — that is, an informed agency, but not necessarily a poorer programmatic outcome. This matters when determining responses to a program. If this pattern was due to access failure, it would be a demand-side intervention. Because it more plausibly reflects women exercising genuine choice once informed, the intervention is instead to extend that same quality of informed choice — full side-effect disclosure, real method variety, and low-friction switching pathways — to the less-educated majority of Nigerian contraceptive users, for whom persistence with a poorly tolerated method may be the less visible but more consequential problem.

### 5.5 What the Algorithm Comparison Itself Contributes

The methodological approach employed in this study (all four algorithms, both outcomes alike, both evaluated using a single, consistently weighted pipeline in Python, and all three models using elastic net, RF, and XGBoost) is a finding in itself. Both times, Random Forest outperformed all other models, which should give it credibility beyond what a single-outcome study could provide. XGBoost, by contrast, went from the second-best algorithm on the larger, more strongly signaled Gap A outcome to the weakest performer on the smaller, more evenly balanced Gap B outcome — underperforming even the simple logistic baseline in Gap B. This is not a modelling error or a contradiction; it is a real, public finding, one which passed a significant methodological correction (the switch from an unweighted linear approximation towards a properly weighted logistic model, plus an omitted predictor) rather than being a modelling error or a contradiction. The flexibility of XGBoost is helpful in cases where a large sample is associated with a strong learnable signal (Gap A: LARC status includes a tremendous amount of predictive power), and hindering when the sample size is small and the signal is relatively low in many of the weak predictors (Gap B: the signal is low in many of the predictors, and the sample size is small). This pattern directly echoes the closest methodological precedent available in this literature — comparative machine-learning applications to Mexico’s ENSANUT survey, where XGBoost’s performance advantage over simpler methods was found to be inconsistent across outcomes and specifications — and adds a second, independent confirmation that the choice of “best” algorithm in DHS-scale survey prediction is empirical and outcome-specific, not a matter that can be settled once and assumed to generalize. The take-away message from this is simple to keep in mind for applied researchers using DHS or other survey-based data on other cascade-type health measures: perform the comparison for each measure instead of assuming the algorithm that worked best for an earlier application, which may have been on a different scale.

### 5.6 Policy and Program Recommendations

The findings are brought together in four concrete, evidence-based recommendations:

1. Report national discontinuation by reason category, instead of restricting to method and region as current DHS-based reporting does, to help program managers to differentiate between the roughly half of discontinuations that are method-related (and hence an indicator of service quality) and the roughly half that are intentional (and hence not a blueprint for service improvement).
2. This study identifies the unmanaged side effects of contraceptive methods, particularly those affecting menstruation (unmanaged in short-acting and even long-acting hormonal methods), as the largest potential addressable share of discontinuation, and the scope of practice of pharmacists and PPMVs is extended to address this directly in this study.
3. This study’s evidence demonstrates that investment in access to LARC methods results in much more effective and persistent coverage than access to short-acting methods alone, not just because of the effectiveness of LARC methods, but because of their dramatically increased resistance to the factors that cause coverage to be lost, and therefore less effective.
4. Invest in formulation and delivery-system research and adoption — the study shows the structural mechanism (reliance on repeated user or facility action) that accounts for the largest proportion of the discontinuation gap between method categories is the technology of the formulation and delivery system, and it is truly an intersection between reproductive health policy and pharmaceutical/biotechnology innovation, not just a common topic.

### 5.7 Limitations

Two predictors identified in the original study design — husband/partner approval of family planning and counseling received at method initiation — are not retrospectively available in DHS calendar data and could not be included, despite both being directly relevant to the quality-of-care mechanisms this discussion emphasizes; their absence likely means this study’s estimate of “method-related, addressable” discontinuation is conservative, since counseling quality is plausibly one of its strongest unmeasured predictors. The Gap B analytic sample (3,701 episodes) is meaningfully smaller than Gap A’s (11,624 episodes), which both widens confidence intervals and required a technical correction (singleunit(centered)) for a small-subpopulation survey design issue; results for Gap B should accordingly be treated as somewhat less stable than Gap A’s, and would benefit from replication as future DHS rounds accumulate more episodes. The categorization of the reasons for discontinuation as “method-related” versus “intentional” is an analytic classification, not an objective classification, and reasons for discontinuation that are construed as “intentional” and not addressed in counseling could be subject to a reader or future replication to reasonably reconsider. Lastly, as in all DHS calendar-based analyses, this study depends on the self-report of all variables with possible recall bias, and predictor variables (education, wealth, insurance) are also measured at the time of interview, and not necessarily at the time of the historical episode, although this was somewhat addressed by the use of the calculated age-at-episode-start, rather than current age.

## 6. Conclusion

The problem of contraceptive discontinuation in Nigeria, as can be seen in this study, is not a single problem, but two, broadly speaking of equal magnitude: a fertility-intention story, which policy should honor as opposed to target, and a service-quality story, which is largely focused on unmanaged side effects, product limitations, and gaps in counseling, which policy, and pharmaceutical and biotechnological innovation in particular, can directly address. This result is very strong for a single-country study because the method-type finding is consistent across four independently specified models: one interpretable regression model and three properly weighted machine learning classification models. This study’s findings do not mean that Nigeria is losing new users of contraceptives to discontinuation; it means that the health system Nigeria has reached is losing users to discontinuation and that this can be addressed by helping the users it has already reached to continue being protected, through better counseling, engaging more with the pharmacists and community health providers, and continued investments in the longer-acting and lower-maintenance contraceptive technologies that this study identified as already succeeding where the shorter-acting methods are failing.

## Declarations

## Ethics Approval and Consent to Participate

The secondary data used for analysis in this study are de-identified and publicly available from the 2023–24 Nigeria Demographic and Health Survey (NDHS). The original survey team obtained informed consent from all interviewees, and the original NDHS protocol was approved by the National Health Research Ethics Committee (NHREC), Nigeria, and the ICF Institutional Review Board (IRB). This analysis relied on de-identified secondary data that is accessible via registered, authorized access to the DHS Program repository, so it was not reviewed for additional institutional review.

## Clinical Trial Registration

N/A

## Artificial Intelligence Statement

This work is not generated by a Generative Artificial Intelligence (G.A.I.) or large language model (L.L.M.) tool. The information provided is the authors’ own views and opinions.

## Acknowledgements

The authors are thankful to Ahead Labs, IIT Roorkee, for providing Stata 15 and the necessary skillset for this project. The authors also acknowledge Marwadi University for providing the resources that helped in the successful conduct of the research.

## Data Availability Statement

Data analyzed in this study are available from the DHS Program (https://dhsprogram.com) but are not publicly available because of restrictions to protect the respondents’ confidentiality. Access is only granted with registration and approval by the DHS Program, upon reasonable request at https://dhsprogram.com/data/available-datasets.cfm. The dataset used was the Individual Recode (women’s) file of the 2023–24 Nigeria Demographic and Health Survey (NDHS). We abstracted additional literature and information from publicly available clinical trial information and World Health Organization (WHO) reports.

## Large-Language Model (LLM)

N/A

